# Interference control and associated brain activity in children with familial high-risk of schizophrenia or bipolar disorder – A Danish register-based study

**DOI:** 10.1101/2022.10.28.22281552

**Authors:** Line Korsgaard Johnsen, Kit Melissa Larsen, Søren Asp Fuglsang, Anna Hester Ver Loren van Themaat, William Frans Christiaan Baaré, Kathrine Skak Madsen, Kristoffer Hougaard Madsen, Nicoline Hemager, Anna Krogh Andreassen, Lotte Veddum, Aja Neergaard Greve, Ayna Baladi Nejad, Birgitte Klee Burton, Maja Gregersen, Heike Eichele, Torben E. Lund, Vibeke Bliksted, Anne Amalie Elgaard Thorup, Ole Mors, Kerstin Jessica Plessen, Merete Nordentoft, Hartwig Roman Siebner

**Affiliations:** Danish Research Centre for Magnetic Resonance, Centre for Functional and Diagnostic Imaging and Research, Copenhagen University Hospital - Amager and Hvidovre, Copenhagen, Denmark; Child and Adolescent Mental Health Centre, Copenhagen University Hospital, Mental Health Services, Capital Region, Denmark; Copenhagen Research Center for Mental Health – CORE, Mental Health Centre Copenhagen, Copenhagen University Hospital - Gentofte, Mental Health Services, Capital Region, Denmark; Institute for Clinical Medicine, Faculty of Health and Medical Sciences, University of Copenhagen, Copenhagen, Denmark; Radiography, Department of Technology, University College Copenhagen, Copenhagen, Denmark; Department of Applied Mathematics and Computer Science, Technical University of Denmark, Lyngby, Denmark; The Lundbeck Foundation Initiative for Integrative Psychiatric Research (iPSYCH), Aarhus, Denmark; Medical & Science, Clinical Drug Development, Novo Nordisk A/S; Department of Clinical Medicine, Faculty of Health and Medical Services, Aarhus University, Aarhus, Denmark; Center of Functionally Integrative Neuroscience, Aarhus University Hospital, Aarhus, Denmark; Department of Child and Adolescent Psychiatry, Copenhagen University Hospital – Psychiatry Region Zealand, Roskilde, Denmark; The Psychosis Research Unit, Aarhus University Hospital, Aarhus, Denmark; Regional resource centre for autism, ADHD and Tourette syndrome Western Norway, Division of Psychiatry, Haukeland University Hospital, Norway; Division of Child and Adolescent Psychiatry, Department of Psychiatry, The University Hospital of Lausanne (CHUV) and University of Lausanne, Switzerland; Department of Neurology, Copenhagen University Hospital Bispebjerg, Copenhagen, Denmark

**Keywords:** Functional magnetic resonance imaging, flanker task, neurocognitive functioning, genetic predisposition, endophenotype, cohort study

## Abstract

**Background and hypotheses:** Impaired interference control is a potential prognostic and endophenotypic marker of schizophrenia (SZ) and bipolar disorder (BP). Assessing children with familial high-risk (FHR) of SZ or BP enables characterization of early risk markers and we hypothesize that they express impaired interference control as well as aberrant brain activation compared to population-based control (PBC) children.

**Study design:** Using a flanker task, we examined interference control together with functional magnetic resonance imaging (fMRI) in 11-to-12-year-old children with FHR of SZ (FHR-SZ) or FHR of BP (FHR-BP) and population-based control (PBC) children as part of a register-based, prospective cohort-study; The Danish High Risk and Resilience study – VIA 11.

**Study results:** We included 85 (44 % female) FHR-SZ, 63 (52 % female) FHR-BP and 98 (50 % female) PBC in the analyses. Interference effects, caused by the spatial visuomotor conflict, showed no differences between groups. Bayesian ANOVA of reaction time (RT) variability, quantified by the coefficient of variation (CV_RT_), revealed a group effect with similarly higher CV_RT_ in FHR-BP and FHR-SZ compared to PBC (BF_10_ = 6.82). The fMRI analyses revealed no evidence for between-group differences in task-related brain activation. Post-hoc analyses excluding children with psychiatric illness yielded same results.

**Conclusion:** FHR-SZ and FHR-BP at age 11-to-12 show intact ability to resolve a spatial visuo-motor conflict and neural efficacy. The increased variability in RT may reflect difficulties in maintaining sustained attention. Since variability in RT was independent of existing psychiatric illness, it may reflect a potential endophenotypic marker of risk.

## Introduction

Schizophrenia (SZ) and bipolar disorder (BP) are severe, highly heritable^1^ psychiatric disorders.^2^ Children of parents with SZ or BP (i.e. familial high-risk; FHR) have twice as high risk for developing a severe mental disorder (SMD) compared to the general population^3^, together with a wide range of other mental disorders before adulthood.^4^ Therefore, investigations in children with FHR of SZ or BP can contribute to the identification of factors of early neurodevelopmental vulnerability. Interference control is a central process of cognitive control^5^ and represents the capability to detect and filter out irrelevant or conflicting information to the task at hand.^6,7^ Effective interference control relies on the integrity of brain systems that mediate information, attention, and inhibitory processing, such as the anterior cingulate cortex (ACC), sub-areas of the parietal cortex, the premotor area, and the insula.^8^ Impaired interference control leads to inappropriate or impulsive behaviors which are common in individuals with neurodevelopmental disorders, including SZ, BP, and attention-deficit/Hyperactivity disorder (ADHD).^9–11^ Altered interference control and correlated brain function have been reported in several studies, using functional magnetic resonance imaging (fMRI) in individuals diagnosed with SZ^12,13^ and BP^12,14^, as well as in individuals at clinical high-risk of SZ^13^ or BP^15^, and adults with FHR^16–23^, emphasizing its potential as a prognostic and endophenotypic marker. Only a limited number of neurobiological investigations of young individuals (age <18) exist but show that young individuals (age 8-25 years) with FHR of SMD display aberrant task-related brain activation in fMRI studies investigating selective attention and/or interference or inhibitory processing.^24–27^ However, sample sizes are small, results between studies are divergent, and studies are of cross-sectional design, impeding finite conclusions.^28^ This motivated The Danish High Risk and Resilience Study that investigates a large prospective cohort of age matched children with and without FHR of SZ or BP.^29,30^

In the first wave of the Danish High Risk and Resilience Study, The VIA 7-study, seven-year-old children with FHR for SZ and BP exhibited behavioral neurocognitive impairments^31–34^ that persisted as a stable developmental deficit at age 11.^35^ Specifically, interference control was impaired along with higher variability in reaction times at age seven in children with FHR of SZ compared to controls.^31^ The impairment in interference control at age seven may constitute an early endophenotype for the later development of SZ and BP^36^, but has not been investigated longitudinally. Further, little is known about the functional properties of the neural networks mediating interference control impairments in children with FHR of SZ or BP during the pre-pubertal neurodevelopmental period.

In the first follow-up of the VIA 7 study, The VIA 11 study^30^, we re-assessed the cohort, now age 11, using the Flanker task in neuroimaging settings of electro-encephalography^37^ and fMRI. For the present paper we use the blood-oxygen level dependent (BOLD) signal obtained with fMRI to probe functional brain activation during interference control in the VIA 11 cohort. We hypothesized that children with FHR-SZ and FHR-BP would be impaired at the behavioral level when confronted with high demands on interference control. Further, we expected that FHR-SZ and FHR-BP children would display aberrant neural engagement of brain regions within the multiple demand network compared with PBC. The flanker-EEG analyses will be presented in a separate paper.

## Methods

Data acquisition was conducted from March 1^st^, 2017 until June 30^th^, 2020 at two sites in Denmark. For a detailed description of data acquisition and analyses procedures see the eMethods.

### Participants

Children were recruited through The Danish High-Risk and Resilience study – VIA 11, including children with at least one parent with SZ (i.e., FHR-SZ) or BP (i.e., FHR-BP), and children with parents without these two disorders (i.e., PBC). The cohort and overall study design are detailed elsewhere^29,30^, and in the eMethods. Written informed consent was obtained from the parent or legal guardian of the child. Children received gift cards for their participation. The VIA 11 study was approved by the National Committee on Health Research Ethics (Protocol number: H 16043682) and the Danish Data Protection Agency (ID number RHP-2017-003, I-suite no. 05333) and conducted in accordance with the Declaration of Helsinki.

### Descriptive clinical measures

Child assessors were blind to FHR status. Axis-I disorders were identified through the Kiddie Schedule for Affective Disorders and Schizophrenia for School-Age Children-Present and Lifetime Version (K-SADS-PL).^38^ The child’s level of functioning was assessed with the Children’s Global Assessment Scale (CGAS)^39^ as part of the K-SADS-PL-interview. The child’s level of emotional and behavioral problems was assessed with the Child Behavior Check List (CBCL) School-Age version.^40^ Handedness was assessed with the Edinburgh Handedness Inventory (EHI).^41^

### Flanker task design

The children completed one ~10-minute session of a modified arrow-version of the Eriksen flanker task^42,43^ (eFigure 2) during fMRI at 3 Tesla to examine interference control and processing.

### Behavioral outcome measures

Interference control was evaluated through response accuracy (resp-acc), mean reaction time (RT), global RT variability (defined as coefficient of variation [CV] through the equation: 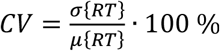, denoted RT_CV_) on congruent and incongruent trials separately, as well as the flanker effect on resp-acc (Δ_resp-acc_), RT (Δ_RT_) and RT_CV_ (Δ_RTCV_).

Additionally, to investigate between-group differences in the speed-accuracy trade-off associated with fast sensorimotor interference tasks, resp-acc (Dist-resp-acc) on congruent and incongruent trials, as well as Δ_resp-acc_(Dist-Δ_resp-acc_) and Δ_RT_(Dist-Δ_RT_), were analyzed with a RT distribution-analytical technique.^44^ This approach allows for the assessment of the temporal dynamics associated with response conflict^44^ and output performance measures (e.g. resp-acc, RT, Δ_resp-acc_, and Δ_RT_) in relation to RT distribution quartiles denoted time-bin 1 through 4.

### Statistical analyses

Statistical tests on behavioral and clinical outcome measures were performed in SPSS (IBM SPSS Statistics version 25, release 25.0.0.2) and JASP (JASP Team (2020), Version 0.8.1).^45^ We used Bayesian ANOVA for testing group differences on handedness, CBCL and CGAS scores. In case of non-distributed data, we used the non-parametric Kruskal-Wallis or Pearson chi-square test with three groups, see Table 1 and eTable 5 for details. Bayesian RM ANOVAs were used for testing group differences on the behavioral outcome measures of RT and CV_RT_. Models included Group as between-subject factor with three levels (FHR-SZ, FHR-BP, and PBC) and Condition as within-subject factor with two levels (congruent, incongruent). Δ_resp-acc_, Δ_RT_ and Δ_CVRT_ were tested for group differences by separate Bayesian ANOVAs with Group as fixed effect.

**Table 1:**
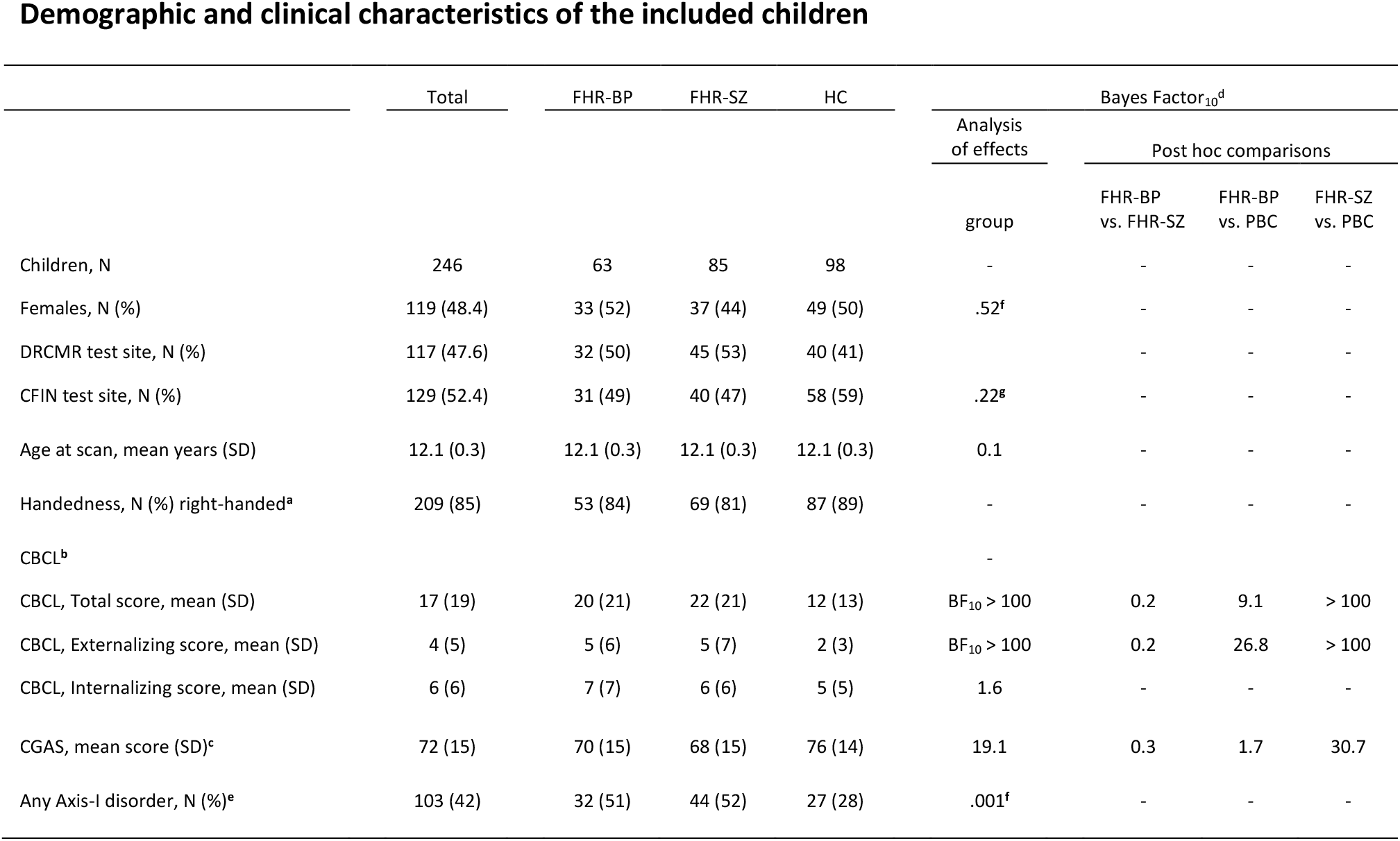
Demographic and clinical characteristics of 84 children at familial high-risk of schizophrenia (FHR-SZ), 63 children at familial high-risk of bipolar disorder (FHR-BP), and 98 population-based control (PBC) children from the Danish High Risk and Resilience Study – VIA 11. DRCMR; Danish Research Center for Magnetic Resonance, CFIN; Center of Functionally Integrative Neuroscience, SD; standard deviation CBCL; Child-behavior check list, CGAS; Child Global Assessment Scale. ^**a**^ Handedness was assessed with the Edinburgh handedness inventory (33) (See Clinical measures) and data is presented as % right-handed according to the laterality quotient score. ^**b**^ Two participants (1.4 %) did not complete the Child Behavior check list (CBCL). Higher scores indicate more problem behavior on the total score as well as on the two broad-band subscales; Internalizing and Externalizing scores. ^**c**^ Higher scores (0-100) on the Child Global Assessment Scale (CGAS) indicate higher global level of functioning. ^**d**^ Bayes Factors (BF) where estimated with Bayesian analyses of variance (ANOVA) in favor of an effect of group (the alternative hypothesis), i.e., BF10, using default priors and 1000 iterations (see Methods). ^**e**^ Quantification of Any Axis-I disorder was obtained with the Schedule for Schedule for Affective Disorders and Schizophrenia for School-Age Children-Present and Lifetime Version (K-SADS-PL) and depicts number of children with any past or present Axis-I diagnosis excluding elimination disorders. ^**f**^ Pearson chi-square test of independence. ^**g**^ Kruskal-Wallis test

|  | Total | FHR-BP | FHR-SZ | HC | Bayes Factor<sub>10</sub><sup>d</sup> |  |  |  |
| --- | --- | --- | --- | --- | --- | --- | --- | --- |
|  |  |  |  |  | Analysis of effects | Post hoc comparisons |  |  |
|  |  |  |  |  |  | FHR-BP vs. FHR-SZ | FHR-BP vs. PBC | FHR-SZ vs. PBC |
| Children, N | 246 | 63 | 85 | 98 | - | - | - | - |
| Females, N (%) | 119 (48.4) | 33 (52) | 37 (44) | 49 (50) | .52<sup>f</sup> | - | - | - |
| DRCMR test site, N (%) | 117 (47.6) | 32 (50) | 45 (53) | 40 (41) |  | - | - | - |
| CFIN test site, N (%) | 129 (52.4) | 31 (49) | 40 (47) | 58 (59) | .22<sup>g</sup> | - | - | - |
| Age at scan, mean years (SD) | 12.1 (0.3) | 12.1 (0.3) | 12.1 (0.3) | 12.1 (0.3) | 0.1 | - | - | - |
| Handedness, N (%) right-handed<sup>a</sup> | 209 (85) | 53 (84) | 69 (81) | 87 (89) | - | - | - | - |
| CBCL<sup>b</sup> |  |  |  |  | - |  |  |  |
| CBCL, Total score, mean (SD) | 17 (19) | 20 (21) | 22 (21) | 12 (13) | BF<sub>10</sub> > 100 | 0.2 | 9.1 | > 100 |
| CBCL, Externalizing score, mean (SD) | 4 (5) | 5 (6) | 5 (7) | 2 (3) | BF<sub>10</sub> > 100 | 0.2 | 26.8 | > 100 |
| CBCL, Internalizing score, mean (SD) | 6 (6) | 7 (7) | 6 (6) | 5 (5) | 1.6 | - | - | - |
| CGAS, mean score (SD)<sup>c</sup> | 72 (15) | 70 (15) | 68 (15) | 76 (14) | 19.1 | 0.3 | 1.7 | 30.7 |
| Any Axis-I disorder, N (%)<sup>e</sup> | 103 (42) | 32 (51) | 44 (52) | 27 (28) | .001<sup>f</sup> | - | - | - |

Resp-acc data for congruent and incongruent trials, as well as in the RT distributional analyses, violated tests for normality and inference on resp-acc was therefore explored using automated non-parametric testing (Kruskall-Wallis) for independent samples in SPSS.

Normal distributed data for the RT distributional analysis was analyzed with separate Bayesian RM ANOVAs including Δ_resp-acc_ and Δ_RT_, respectively, as RM factor with four levels (RT bin 1-4). Group was entered as between-subject factor with three levels (FHR-BP, FHR-SZ, HC). Age, sex, and test-site were entered as covariates in all analyses. If evidence for effects of single covariates were present in the Bayesian analyses, analyses were rerun including covariates as between-subject factors for inference on group by covariate effects. Group effects that met evidence for the alternative hypothesis larger than moderate, were tested with post-hoc comparisons with null control.

fMRI data was analyzed using a general linear model (GLM) in SPM12 (Wellcome Centre for Human Neuroimaging) run in MATLAB (The MathWorks, Inc., version R2020a update 3). First-level (i.e., subject-level) analyses included task-relevant regressors of response hand (right or left), condition (congruent or incongruent), errors (commission or omission) and feedback stimuli presentation and task-irrelevant regressors of movement and physiological noise (See eMethods for complete report). An incongruent>congruent contrast was formed based on task regressors and used to model successful interference activation as we only included trials in which the participants answered correctly. At the group-level, the main contrasts were analyzed in a univariate flexible factorial ANOVA with group (FHR-BP, FHR-SZ, PBC) as between-subject factor. We applied a cluster-forming threshold of *P* < .001, uncorrected, and a Gaussian random field-based family wise error (FWE) correction of *P* < .05 at the cluster level for reporting of significant clusters of brain activation and for defining peak activation coordinates for centers of regions of interest (ROI). For quantification of brain activity in linear relation to RT distribution, we modelled fMRI data according to the RT bins by adding the RT bin quartiles as a first parametric modulator of interest in the first-level analyses. Brain-behavior correlations were performed on behavioral variables showing group effects. The fMRI analyses included age, sex, handedness, and test-site as covariates.

Additionally, we analyzed mean parameter estimates from the contrasts in ROI analyses within the Bayesian framework to characterize evidence for or against the alternative hypothesis. ROIs were defined as 10 mm spheres with their center placed according to peak activation coordinates (cluster-level FWE corrected *P*<.05) from the main effect of the successful interference activation analysis across groups. ROIs were included as repeated measures (RM) factor with nine levels (ROI I, ROI II, etc.) and group as between subject factors with three levels (i.e. FHR-BP, FHR-SZ, PBC) in a Bayesian RM ANOVA implemented in JASP.^45^

## Results

Demographic and clinical characteristics of included children are presented in Table 1. The final sample consisted of 246 children: 85 children (44 % female) with FHR-SZ, 63 children (52 % female) with FHR-BP and 98 (50 % female) PBC. A full overview and description of the inclusion procedure for the present analyses can be seen in eFigure 1 and is further detailed in the eMethods. The three groups were comparable on age (BF_10_=0.1, moderate evidence against a group effect), handedness (BF_10_=0.09, strong evidence against group effect), and sex (Pearson chi-square, *P*=.52). Children with FHR-SZ and FHR-BP presented with higher behavioral problem scores compared to PBC, and lower global functioning scores for FHR-SZ compared to PBC. There was higher incidence of Axis-I disorders in the FHR groups (Pearson chi-square, *P*<.001).

**Figure 1:**
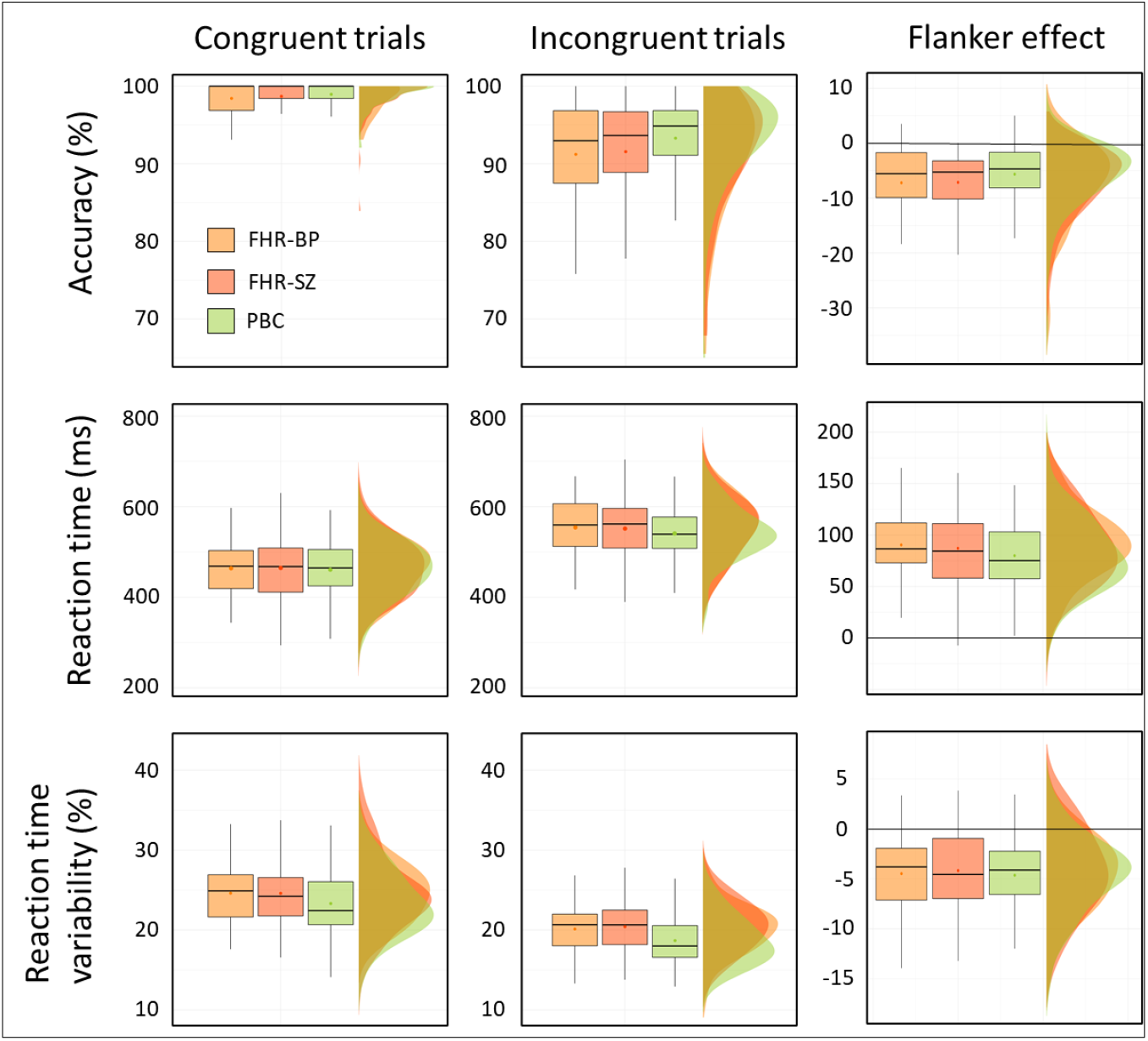
Task performance measures, i.e., response accuracy, reaction time and reaction time variability on the modified arrow-version of the Eriksen flanker task for children at familial high-risk of schizophrenia (FHR-SZ; dark orange), bipolar disorder (FHR-BP; light orange) or neither of these disorders (Population-based controls [PBC]; green). Data has been plotted as typical boxplots illustrating the minimum, maximum, median, first quartile and third quartile, as well as the data distribution clouds to match. The dot within each boxplot illustrates the mean value. Performance across the three groups shows an overall decrease in accuracy rate (top horizontal panel), and an overall increase in reaction time (middle horizontal panel), from congruent (left vertical panel) to incongruent (middle vertical panel) trial conditions as well as the difference (i.e., the flanker effect) in these measures (right vertical panel). Values larger than zero indicate an increase in performance measure from incongruent to congruent conditions whereas values lower than zero indicate a lower performance measure. The bottom horizontal panel shows reaction time variability, calculated as the coefficient of variation 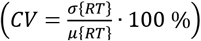 in which there was moderate evidence (Bayes factor [BF]_10_ = 6.8) for an effect of group but moderate evidence against (BF_10_ = 0.22) a group by condition (i.e., congruent and incongruent) interaction. ms; milliseconds, FHR-SZ; Familial high-risk of schizophrenia, FHR-BP; Familial high-risk of bipolar disorder, PBC; Population-based controls.

### Behavioral interference control

The three groups performed similarly on resp-acc across conditions (*P*=.218, congruent, *P*=.135, incongruent), with all groups showing higher accuracies on congruent trials compared to incongruent trials (Figure 1, top row, and supplemental eTable 5). All groups showed slower RT for incongruent trials compared to congruent trials (BF_10_>100) with similar mean values (BF_10_=.22), see Figure 1, middle row.

Across the two conditions the variability of response timing RT_CV_ differed between groups (BF_10_=6.88) indicating that children with FHR showed higher RT_CV_ compared to PBC, Figure 1 bottom row. Sex and test-site showed strong (BF_10_=19.9) and decisive (BF_10_>100) effects on RT_CV_, respectively. Adding these as between-subject factors revealed that these did not interact with the group effect.

As expected, the speed-accuracy trade-off analysis revealed an effect of time-bin for the congruent (Friedman’s ANOVA, *P*=.034) and incongruent (Friedman’s ANOVA, *P*<.001) condition, showing higher accuracies in the slowest trials. This trade-off was similar across all three groups, Figure 2B. Groups did not differ on the Flanker effect for either accuracy (Dist-Δ_resp-acc_ BF_10_=.11) or RT (Dist-Δ_RT_ BF_10_=.15), Figure C and D, respectively. For a summary of the statistical output see eTable 6.

**Figure 2:**
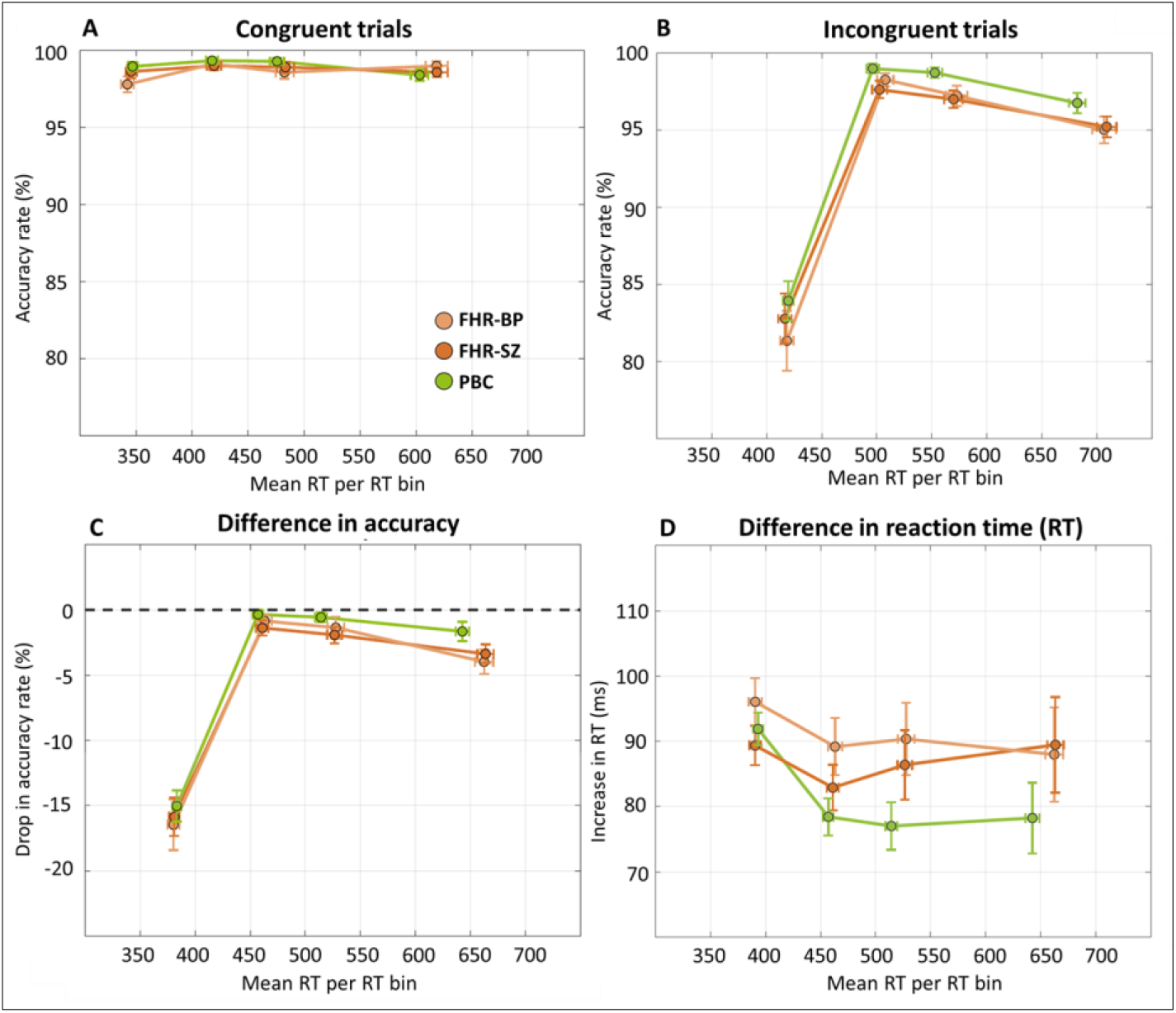
Reaction time (RT) distribution-analysis of response accuracy rate on congruent (A) and incongruent (B) trials (Dist-resp-acc). Delta plots show the magnitude of interference effects on response accuracy rate (C; Dist-Δ_resp-acc_) and RT (D; Dist-Δ_RT_) as a function of RT on the modified arrow-version of the Eriksen flanker task for children at familial high-risk of schizophrenia (FHR-SZ), bipolar disorder (FHR-SZ) or neither of these disorders (Population-based controls; PBC). Mean RTs are plotted according to the 1st, 2nd, 3rd, and 4th quartile of RTs according individual RT distributions for correct-proceeded-by-correct trials (See supplemental methods). Error bars indicate standard error of the mean (SEM) calculated by 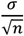 and illustrated for both mean RTs of the quartiles as well as the accuracy rates (A-C) and the drop in RT (D).

### Interference-related brain activation

To ensure that our task engaged a relevant interference network, we investigated the main congruency effect on brain activity. Across groups we found significant activation in nine clusters (Figure 3A, supplementary eTable 2); left cerebellar cruz II, inferior division of right and left lateral occipital cortex, left and right insula, right precentral gyrus, left supplementary motor area (SMA), right middle frontal gyrus (MFG), and left superior frontal gyrus (all p < 0.05, cluster-level corrected). We found no significant main effect of group in the whole-brain analyses regarding successful interference control. This was confirmed in the ROI analysis using Bayesian inference (Figure 3B, eTable 6 A).

**Figure 3:**
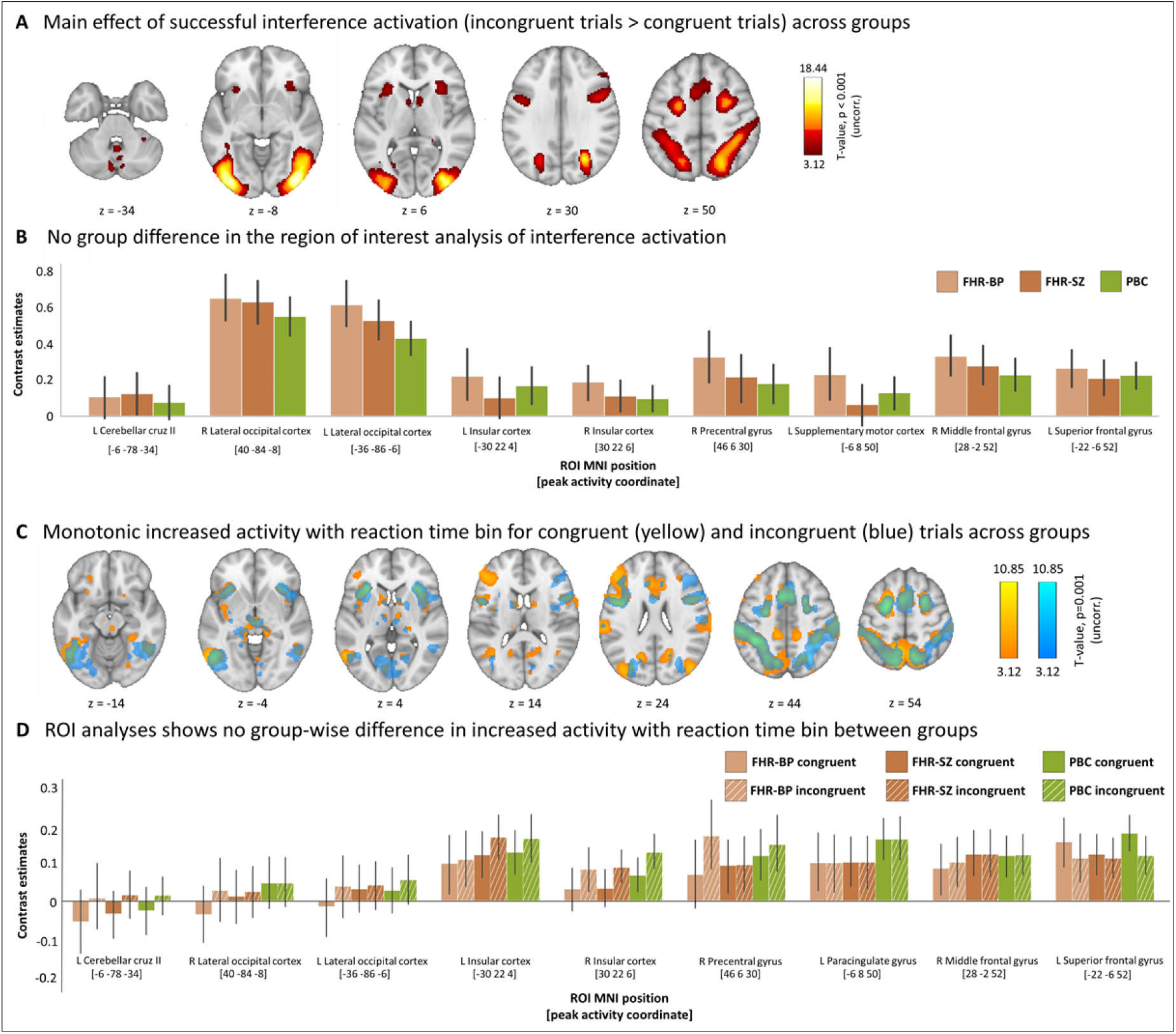
Interference control related brain activation results from 63 children with familial high-risk (FHR) of bipolar disorder (FHR-BP), 85 children with FHR of schizophrenia (FHR-SZ) and 98 population-based control children without FHR of these disorders. (A) The group-level whole-brain analysis on the contrast between correct incongruent trials larger than correct congruent trials (successful interference activation) showed significant increased activation in nine task-related clusters (cluster-level FWE-corrected P < .05) within areas of cerebellum, Left and right lateral occipital cortex, left and right insular cortex, right precentral cortex, left supplemental motor area, middle frontal gyrus, and superior frontal gyrus. The color bar indicates T-values. The analysis did not show any significant group differences or any significant condition by group interactions. (B) Bar plot showing contrast estimates (CE) from the region of interest (ROI) analysis on the successful interference activation. Error bars indicate the standard deviation. (C) Whole-brain activation modulated by reaction time (RT) bin on correct congruent trials larger than 0 (yellow) and correct incongruent trials larger than zero (blue) across groups. There was no significant between group difference. (D) Bar plot showing contrast estimates (CE) from the region of interest (ROI) analysis on the brain activation modulated by RT bin analysis. Congruent contrast estimates are solid bars, incongruent contrast estimated are scratched bars. Error bars indicate the standard deviation. Incongruent contrast estimates are shown with hatched bars.

The speed-accuracy trade off analysis including time-bin as a first parametric modulation of interest revealed 10 significant clusters of activation spanning insular, fronto-temporal, striatal, thalamic, and cingulate brain areas for the congruent condition (Figure 3C and eTable 3). For the incongruent condition nine significant clusters of activation were located within temporo-parietal, thalamic, striatal, and occipital brain areas (Figure 3C and eTable 3). This network was stable across groups and confirmed in a follow up ROI analysis using Bayesian inference (Figure 3D, and eTable 6 B and C).

To follow-up on the behavioral findings of moderate evidence for an effect of group on RT variability, RT_CV_ was added as a regressor of interest in two independent second level analyses. We entered RT_CV_ for congruent and incongruent trials and used the contrast of congruent > 0 and incongruent > 0 from the first level, respectively. For the congruent contrast, we found a significant negative relationship between RT_CV_ and brain activity in four clusters within occipital and parietal brain areas across the three groups (Figure 4, supplementary eTable 4). This network was stable across groups.

**Figure 4:**
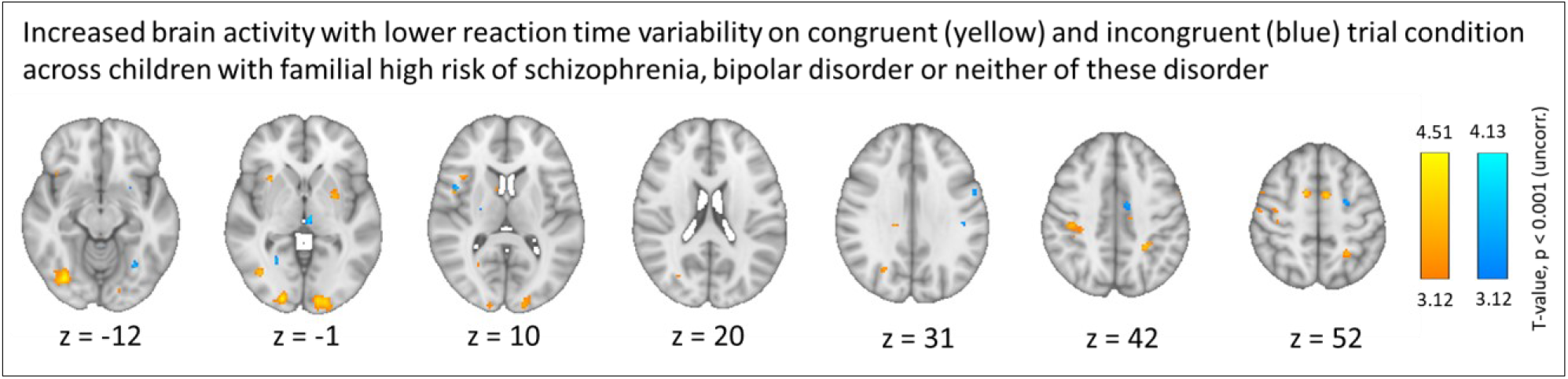
Whole brain blood-oxygen level-dependent (BOLD) response with reaction time (RT) variability – calculated as the coefficient of variation (CV) – as linear regressor of interest across 63 children at familial high-risk (FHR) of bipolar disorder (FHR-BP), 85 children with FHR of schizophrenia (FHR-SZ) and 98 children without FHR of these disorders (population-based controls). Contrast images are based on correct responses on congruent trial events > 0 (yellow) and incongruent trials events > 0 (blue) and show brain activation as a function of lower reaction variability – a reaction time stability network that is.

To disentangle effects of FHR from that of existing psychiatric diagnoses, we replicated all analyses excluding all children with any past or present Axis-I diagnosis (n=103). These post hoc analyses resulted in similar findings as the ones reported in the total sample.

## Discussion

We show evidence that children with FHR for SZ or BP express higher variability in reaction time (RT) during a visuo-motor response conflict task, probing interference control. This variability persisted when excluding children with any past or present psychiatric diagnosis. We therefore argue that this increased variability in response timing might reflect an endophenotypic trait for risk of SZ and BP. The behavioral fingerprint was not accompanied by altered group-wise brain responses, indicating a similar brain network engagement when solving the flanker task.

The increased RT variability, indexed by a higher RT_CV_, was observed in children with FHR-SZ and FHR-BP independently of interference level, showing that the increased variability of RT was independent of the presence or absence of a visuospatial response conflict. A more variable timing of responses in the FHR groups agrees with previous results from this cohort, observed at age seven.^31,32^. It also extends earlier findings in children with FHR of SZ or BP beyond those already reported in this cohort across different paradigms, testing attentional and interference control abilities. ^21,27,46^ Indeed, larger variability of RT is shared across many disorders such as ADHD, SZ and BP^47–49^ and refers to an increased within-subject fluctuation of trial-to-trial behavior making it an attentional attribute related to the maintenance of continuous performance.^50^

In addition to the increased variability in RT at age 7, the children with FHR in the VIA-7 study showed impaired interference control (i.e., response accuracy (resp-acc) and mean reaction time (RT))^31^ which was hypothesized to still be present at age 11. Indeed, separate analyses of 23 neurocognitive measures in this cohort, spanning multiple domains from age 7 to 11 showed a stable developmental deficit for the children with FHR of SZ compared to controls.^35^ We found evidence against between-group differences in interference control at age 11-to-12. This finding suggests that impaired interference control may be of transitory character, or it may be attributed to methodological factors. The flanker paradigm used at age 7 was run at a faster pace with relatively short inter-trial-interval (ITI; 900 ms) and applied in a larger sample size (n=492). In our fMRI study, we implemented a slower version of the task to accommodate the sluggishness of the hemodynamic response. The longer ITIs may have resulted in a less demanding paradigm. It is thus possible that the high-risk groups would have maintained impaired performance in a more challenging setting with shorter ITIs.

Meta-analytical evidence, based on samples with broad age ranges that may obscure neurocognitive developmental factors, have shown impaired interference control using Stroop tasks in young (age 15-29) individuals with FHR of SZ^51^ with small to medium effect sizes (Cohen’s d=0.28 [95% CI 0.04-0.52]), and in youth (mean age 10-25) with FHR of BP^52^ with similar effect sizes (Cohen’s d=0.3 [95% CI 0.11-0.48]) when compared to controls. The Stroop task is, like the Flanker task, used to assess inhibitory abilities of cognitive interference.^53,54^ However, intact interference control measured with Stroop in child and adolescent offspring of patients diagnosed with schizophrenia or bipolar disorder at baseline (age: 10-12 years) and follow-up (age: 12-14 years) have also been shown^55^, leaving this cognitive measure yet unresolved as a candidate endophenotype in young FHR populations.

Our task reliably evoked interference-related brain activation in areas typically associated with interference control, i.e., cerebellar, occipital, parietal, premotor, and prefrontal areas.^8,56–60^ Within this network, task-related brain activation was comparable between our three groups. In child FHR populations, fMRI investigations of task-related activation together with interference control are scarce.^19,28^ To the best of our knowledge, this is the first task-based fMRI study on interference control in children with FHR of SZ. Tapping into inhibitory control, a small fMRI study examined task-related activation during a stop-signal task in 13 adolescents with FHR of BP (mean age ~13 years).^24^ That study reported significantly greater activation in putamen during unsuccessful motor inhibition compared to children without FHR (n=24, mean age ~14).^24^ Another study on cognitive flexibility (using a change task), reported that youth with FHR of BP (n=13, mean age ~14 years) showed increased activity in ventrolateral prefrontal and inferior parietal areas during successful cognitive flexibility and increased brain activation in the caudate nucleus during failed change trials compared to controls (n=21, mean age ~14).^26^ Together these studies point to hyper-activity of dorso-striatal structures as a mechanism for failing inhibitory processes in children with FHR of BP. However, the sample sizes of these two studies are very small, bearing a considerable risk of false positive findings. In addition, the focus on unsuccessful inhibition makes comparison to the current investigation challenging. In our setup, most of the children completed the task without mistakes. This behavior limits us from assessing unsuccessful interference.

Our study has strengths and weaknesses. The present study is the first to include a relatively large number of children (>200) with a similar young age of 11 to 12 resulting in large statistical power. Throughout adolescence, interference control continues to improve until full maturation in adulthood.^61–63^ Therefore, the mean age of the children in our cohort represents a developmentally sensitive time during which continuous brain changes in relation to maturation and behavioral performance occur in parallel.^64^ Using a sample with a narrow age range reduces the risk of obscuring age effects of neurocognitive and brain maturation in the event of a developmental lag.^65^ However, the pre-adolescent age-group may also leave our analysis too early in development to detect neurobiological differences that may arise later in life. The cross-sectional design of the present study, however, precludes any causal interpretations regarding risk of later development of BP and SZ. Longitudinal studies investigating the developmental trajectories of possible cognitive and/or neurobiological deficits are warranted to assess the state or trait nature of these (possible) early endophenotypic markers for SZ and BP. Our follow-up study, The VIA 15 study, will enable us to explore these aspects.

In conclusion, we found no differences in interference control or related brain activity in this large cohort of 11-12-year-old children with FHR-SZ and FHR-BP compared to PBC. However, a higher variability of the timing of responses across trials was observed in the FHR groups, independently of interference effects. This timing variability of responses during the task was also independent of past or present psychopathology, making this behavioral feature a possible endophenotypic trait marker for risk of schizophrenia or bipolar disorder. Longitudinal designs are warranted to illuminate developmental trajectories in individuals that may go on to develop SZ or BP.

## Data Availability

The data that support the findings of this study are not available due to Danish law.

## Acknowledgements

We thank all the children and their families who participated in the Danish High Risk and Resilience Study. A special thanks to Benthe E. Vink, Jonas S. Ingerslev, Daban Sulaiman, Gøkze M. Akkas, Natascha Larsen, Malte Lundby, Anna K. Møller and Agnete H. Albertsen at DRCMR, as well as Christina B. Knudsen, Nanna L. Steffensen, Merete Birk, Anette F. Bundgaard, Oskar Jefsen and Henriette Stadsgaard at CFIN for their effort in data acquisition. We also acknowledge Line Carmichael for coordinating recruitment, Jessica Ohland for handling the database, and Simon Y. Jensen for support in data analysis.

## Notes

### Competing Interest Statement

Hartwig R. Siebner has received honoraria as speaker from Sanofi Genzyme, Denmark, Lundbeck AS, Denmark, and Novartis, Denmark, as consultant from Sanofi Genzyme, Denmark, Lophora, Denmark, and Lundbeck AS, Denmark, and as editor-in-chief (Neuroimage Clinical) and senior editor (NeuroImage) from Elsevier Publishers, Amsterdam, The Netherlands. He has received royalties as book editor from Springer Publishers, Stuttgart, Germany and from Gyldendal Publishers, Copenhagen, Denmark.
During preparation of the manuscript, Ayna Baladi Nejad changed employment to Novo Nordisk A/S.

### Funding Statement

This study was funded by The Lundbeck Foundation Initiative for Integrative Psychiatric Research - iPSYCH (grant number R248-2017-2003) and Innovation Fund Denmark (grant number 6152-00002B). LKJ was funded by The Independent Research Fund (grant number DFF - 6110-00139), Denmark, and KML received funding from Mental Health services - Capital Region of Denmark (no grant number given) as well as the Lundbeck foundation (grant number R322-2019-2311). Hartwig R. Siebner holds a 5-year professorship in precision medicine at the Faculty of Health Sciences and Medicine, University of Copenhagen which is sponsored by the Lundbeck Foundation (grant number R186-2015-2138). The funders had no role in study design, data collection, data analysis, data interpretation, decision to publish or preparation of the manuscript.

### Author Declarations

Ethics commmittee/IRB of the National Committee on Health Research Ethics gave ethical approcal for this work (Protocol number: H 16043682)

