## Supplemental methods, figures and tables for "Interference control and associated brain activity in children with familial high-risk of schizophrenia or bipolar disorder – A Danish register-based study"

**Inclusion of participants**

The original cohort in The Danish High Risk and Resilience Study - VIA 7 was established from 2013 to 2016, and consist of children with at least one parent diagnosed with SZ (FHR-SZ, n = 202) or BP (FHR-BP n = 120), and children with parents without any of these two disorders (population-based control; PBC, n = 200). Schizophrenia (SZ) was defined as ICD-10 codes F20, F22, and F25, or ICD-8 codes 295, 297, 298.29, 298.39, 298.89, and 298.99. Bipolar disorder (BP) was defined as ICD-10 codes F30 and F31, or ICD-8 codes 296.19 and 296.39. The children were recruited from the Danish Psychiatric Central Research Register (1) and the Danish Civil Registration System (2). Permission to retrieve the cohort from the Danish national registers was granted by the Danish Ministry of Health. PBC children were matched to the FHR-SZ children on age, sex and municipality (3, 4). In the second wave, the VIA11 study, children with FHR-BP did not differ from the two other groups on these characteristics even though they were a non-matched sample.

The present study focused solely on participants from the VIA 11 Study, investigating the cohort (FHR-SZ n = 179, FHR-BP n = 105, PBC n = 181) at age 11. Of the children participating in the VIA 11 Study, 397 children (FHR-SZ n = 148, FHR-BP n = 91, PBC n = 158) said yes to participate in the MRI session. The full overview of the inclusion/exclusion procedure is shown in eFigure 1. Children underwent fMRI assessment at one of two sites in Denmark; 1) the Danish Research Centre for Magnetic Resonance (DRCMR), Centre for Functional and Diagnostic Imaging and Research, Copenhagen University Hospital – Amager and Hvidovre; 2) the Centre of Functionally Integrative Neuroscience (CFIN), Aarhus University Hospital, Skejby.

On the day of scanning 55 children (FHR-SZ n=11, FHR-BP n=23, PBC n=21) opted out before all relevant data for the present analyses were collected (i.e., functional and structural images) leaving 325 complete datasets. Of these, a total of 79 datasets were not eligible for data analyses due to incidental clinical findings on the structural MR-images or neurological disorder (FHR-SZ n=2, FHR-BP n =1, PBC n=2), software/hardware malfunction during the scan session (FHR-SZ n=10, FHR-BP n=3, PBC n=14), poor data quality due to braces or participant motion (FHR-SZ n=6, FHR-BP n=7, PBC n=2), insufficient performance on the task (i.e., performance accuracy < 70%; FHR-SZ n=10, FHR-BP n=2, PBC n=6), and lastly excess movement during fMRI leading to removal of >20% of volumes (FHR-SZ n=4, FHR-BP n=3, PBC n=7).

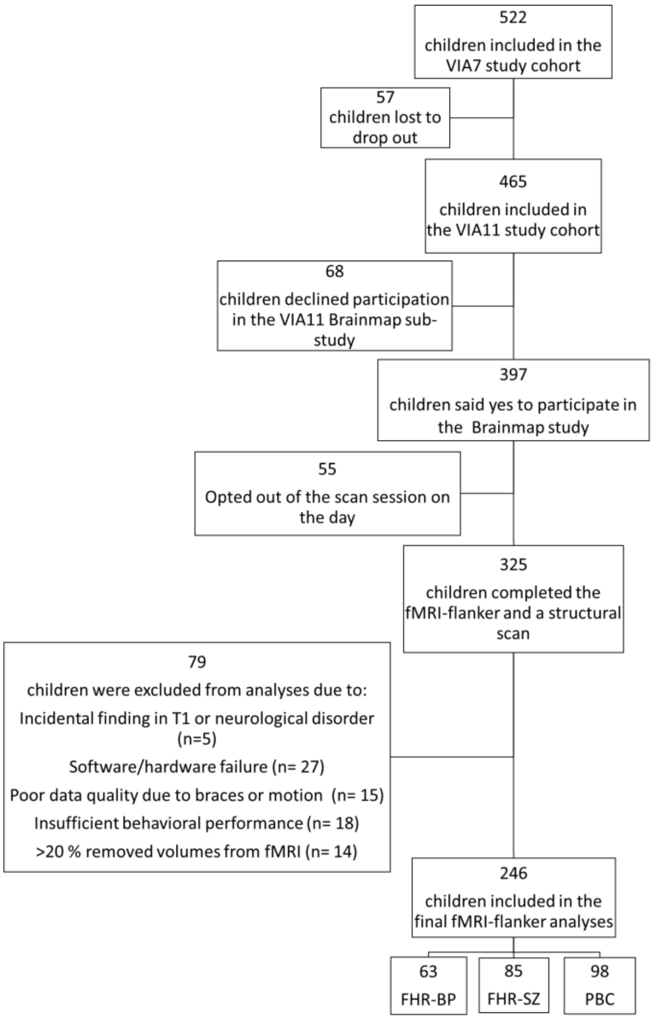

*eFigure 1: Flowchart of the inclusion procedure. FHR-BP; Familial high-risk for bipolar disorder, FHR-SZ; Familial high-risk for schizophrenia, PBC; Population-based controls without any parents with SZ or BP. See supplemental eMethods for specific numbers exclude excluded for each group.*

**Clinical Measures**

Presence of any past or present Axis-I diagnoses was based on the semi-structured diagnostic interview Schedule for Affective Disorders and Schizophrenia for School-Age Children-Present and Lifetime Version (K-SADS-PL). The interview was administered by trained personnel with the child and the primary caregiver. Diagnoses were based on the Diagnostic and Statistical Manual of Mental Disorders (DSM)-IV and DSM-5 Axis-I disorders. Details on the Axis-I diagnoses present in this cohort have been published elsewhere (5). The children's level of behavioral problems was assessed with the Child Behavior Check List (CBCL), School-Age version (6). For CBCL we used the two broad-band subscales (Internalizing and Externalizing) as well as the total score as clinical measures of the children's problem behavior. The questionnaire was completed by the primary caregiver. The primary caregiver was carefully chosen to be the adult knowing the child the best and was, preferably, living with the child. Higher scores on CBCL indicate more problem behavior. Global functioning was assessed with the Children's Global Assessment Scale (CGAS, (7)). Higher scores on CGAS indicate higher global levels of functioning. The CGAS was completed as part of the K-SADS-PL by trained researchers interviewing the child and the primary caregiver. Handedness was determined with the Edinburgh Handedness Inventory (EHI) (8) and characterized as right-handed, left-handed, or ambidextrous according to the obtained laterality quotient (LQ) score; -100 to -50, -50 to 50 and 50 to 100, respectively.

**The flanker practice session**

Before fMRI scanning commenced, children practiced the modified arrow-version of the flanker task inside the scanner. The practice session entailed 10 congruent and 10 incongruent trials, with a between-subjects fixed pseudo-randomized order. Behavioral outcome measures from the flanker training included reaction time (RT), the standard deviation (SD) hereof, and accuracy rate on all trials, excluding omission trials. These variables were used to assess whether children understood and could perform the task, and for determining the feedback stimulus presentation threshold in the subsequent fMRI-flanker task (See **The flanker task design**). A minimum accuracy rate of 70% on incongruent trials was considered to signify a successful training session.

**The flanker task design**

At both scanning sites, the modified arrow-version of the Eriksen flanker task was presented via E-prime software (E-prime 2.0, version 2.0.10.356, copyright 1996-2015 Psychology software tools) and consisted of 128 trials evenly balanced between congruent and incongruent conditions, intermingled with 40 null trials. The trial order was pseudo randomized but fixed between participants. A trial commenced by presentation of six flanking arrows (100 ms), after which a target arrow was presented (1000 ms; response window) in the center of the screen. The target arrow and the flanking arrows faced the same (in congruent trials; low interference-level) or opposing (in incongruent trials; high interference-level) directions. Children were instructed to respond as fast and accurately as possible with their right or left thumb by pressing the response button (Current Designs, 4-Button Curve Right pad on the 8-Button Bimanual Fiber Optic Response Pad) depending on the direction of the target arrow. Right- and left- hand responses were evenly balanced for each condition. Subsequently, a fixation dot or a feedback exclamation mark (!) was presented for 200 ms. The exclamation mark was presented if the recorded RT of the previous trial was greater than the mean RT + 1.5*SD obtained from the flanker training session completed before scanning, or if no response was given within the response window. The children were instructed to speed up their responses if they saw the feedback exclamation mark while also attending to the accuracy of the responses. The feedback exclamation mark was designed to ensure engagement throughout the task. The inter-trial interval was jittered between 1700-1900 ms. On average the trial duration was approximately 3.3 seconds. eFigure 2 shows a schematic of the task structure.

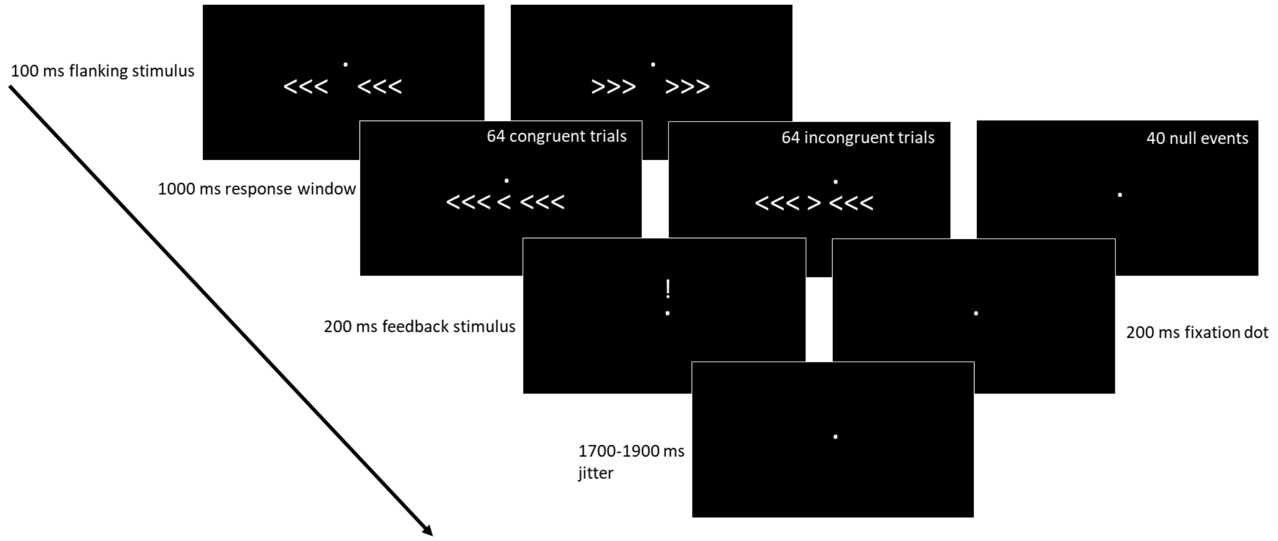

*eFigure 2: Illustration of the modified arrow-version of the Eriksen flanker task. A trial commenced with presentation of the flanking stimulus for 100 ms, followed by a 1000 ms response window, containing either a congruent (64 trials of either <<< < <<< or >>> > >>>), incongruent (64 trials of either <<< > <<< or >>> < >>>) or null events (40 trials with a fixation dot). Congruent and incongruent trials were evenly balanced with right- and left-hand responses. Trials were presented in a pseudo randomized order, fixed between subjects. The feedback stimulus was presented only on occasions where the recorded RT was lower than the mean RT (+ 1.5*SD) obtained at the practice session completed before the scanning session. The inter trial interval was jittered by 1700-1900 ms of fixation dot. ms; milliseconds, RT; reaction time, meanRT; mean reaction time obtained from a flanker training session.*

**Behavioral outcome measures, data analysis and statistical inference**

Behavioral outcome measures are based on the 64 incongruent and 64 congruent trials. The 40 null events were not considered in the behavioral analyses and were removed prior to any processing. Individual trials were removed prior to analysis when 1) the response was given within 200 ms of target onset (considered an anticipatory response), and/or when no response was given within the response window (omission error). This resulted in an average elimination of 4.6% of trials per subject. Using predetermined criteria, children with more than 30% commission errors and/or a response accuracy (resp-acc) on incongruent trials 3*SD below the group mean would be excluded, as we assumed these children to be inadequately engaged with the task. No children were excluded based on these criteria.

For each condition, resp-acc was determined by the number of correct trials divided by the number of total trials (excluding anticipatory trials and omission trials). Responses were defined to be correct if response direction and target arrow corresponded and RT fell within a time window from 200 ms to 1000 ms post target arrow presentation. RT was recorded as the time between target arrow presentation and the first response by button press. $RT_{CV}$ was calculated by dividing the standard deviation (SD) by the mean RT of each participant $\left(CV = \frac{\sigma\{RT\}}{\mu\{RT\}} \cdot 100\ \%\right)$. The $RT_{CV}$ indicates the relative dispersion of RTs around the Mean; hence, it is a measure of RT variability. We use $RT_{CV}$ for easier comparison of performance to the previous report on interference control in this cohort at age seven.(9) Further, using the $RT_{CV}$ as a measure of variability, and not the traditional standard deviation (SD), results in a measure that accounts for mean RT since these have been shown to be highly correlated.(10)

The flanker effect on resp-acc ($\Delta_{resp\text{-}acc}$) and RT ($\Delta_{RT}$) was calculated by subtracting mean resp-acc and mean RT, respectively, on incongruent conditions from congruent conditions. The flanker effect quantifies the difference in the performance measure from incongruent to congruent trials and relates to the increased need for processing of a spatial response conflict that is induced when target and distractors are incongruent.(11)

For the RT distributional analyses, we quantified RT distributions on the subject level, estimating RT-bin thresholds ($25^{th}$, $50^{th}$, $75^{th}$ percentile)(12, 13) from RTs on correct followed by correct trials only, leaving out effects of post-error slowing (14). Hereafter, all trials were assigned to one of the four quartiles and analyzed according to accuracy rate for the congruent and the incongruent condition separately, as well as the flanker effect on accuracy (Dist- $\Delta_{resp\text{-}acc}$) and reaction time (Dist- $\Delta_{RT}$).

*Bayesian inference*

We used uninformed prior odds with r scale fixed effects of 0.5 for all Bayesian statistical models. Levels of evidence are reported according to standard interpretation of Bayes factors (BF) in favor of the alternative hypothesis ($BF_{10}$) ranging from 1 (no evidence) to 1-3 (anecdotal evidence), 3-10 (moderate evidence), 10-30 (strong evidence), 30-100 (very strong evidence), and >100 (*decisive evidence*). (15, 16) The BF quantifies the relative predictive performance of two rival hypotheses.(15)

**Participant preparation**

At DRCMR we used an MRI simulator™ system (Model: PST-100355, Psychology Software Tools Inc.) to introduce the children to the MR scanner environment, with the parent/guardian present. We used the MRI simulator for all participants included at DRCMR. The children were encouraged to lay down on the scanner bed and be moved inside the mock scanner. After some familiarization time, participants were presented to the scanner sounds associated with the different sequences through the mock scanner speakers. The mock-scanner familiarization was not possible at CFIN, but since testers at this site had a longer history with the child (testers at CFIN also completed clinical interviews with the children, whereas testers at DRCMR only performed MRI), the close connection between tester and child at CFIN, was also advantageous at the day of MRI.

In case participants used glasses, they were provided with MR compatible glasses with the right strength.

**MRI acquisition and preprocessing**

Sequence parameters are detailed in eTable 1. Anatomical and functional MR images were acquired in a single session (3.0 Tesla Siemens Magnetom Prisma with a 64-channel head coil [DRCMR]; 3.0 Tesla Siemens Magnetom Skyra with a 32 -channel head coil [CFIN]) using a consistent protocol across sites (eTable 1). Scanning duration for the present analysis was approximately 22 minutes. Total scan time was approximately 01:15 (HH:MM).

| Sequence type | TR (MS) | TE (MS) | TI1 (MS) | TI2 (MS) | Flip angle (degrees) | Voxel size (mm) | Number of slices | Phase encoding direction | Slice orientation | FOV | MB factor | Number of volumes | TA (MM:SS) |
|---|---|---|---|---|---|---|---|---|---|---|---|---|---|
| **DRCMR** | | | | | | | | | | | | | |
| Gradient EPI (fmri-flanker) | 1052 | 30 | - | - | 65 | 2.5 | 54 | A >> P | Transversal | 192 | 3 | 520 | 09:23 |
| Gradient EPI(rev-phase) | 1052 | 30 | - | - | 65 | 2.5 | 54 | A >>P | Transversal | 192 | 3 | 5 | 00:21 |
| MP2RAGE | 6500 | 3.49 | 700 | 2800 | 4 | 0.9 | 192 | A >> P | Sagittal | 260 | - | - | 12:39 |
| **CFIN** | | | | | | | | | | | | | |
| Gradient EPI (fmri-flanker) | 1081 | 30 | - | - | 65 | 2.5 | 54 | A >> P | Transversal | 192 | 3 | 501 | 09:23 |
| Gradient EPI (rev-phase) | 1081 | 30 | - | - | 65 | 2.5 | 54 | A >> P | Transversal | 192 | 3 | 5 | 00:21 |
| MP2RAGE | 6500 | 3.46 | 700 | 2800 | 4 | 0.9 | 192 | A >> P | Sagittal | 260 | - | - | 12:39 |

*eTable 1: MRI sequence parameters applied at DRCMR and CFIN. TR; Repetition time, TE; Echo time, TI; Inversion time, FOV; Field of view, MB; Multi band, TA; Acquisition time, EPI; Echo planar imaging, MPRAGE; magnetization-prepared rapid gradient echo MP(2)RAGE; Magnetization-prepared (2) rapid gradient echo.*

*Study specific template*

From MP2RAGE images, we created a study specific anatomical study template using the DARTEL nonlinear image registration procedure (17) implemented in Statistical Parametric Mapping (SPM) 12 (Version: 12.7771, Wellcome Department of Cognitive Neurology, Institute of Neurology, London, UK; http:// www.fil.ion.ucl.ac.uk/spm) running in Matlab 2020a (The MathWorks, Natick, MA).

*fMRI preprocessing details*

In a separate sequence, Gradient EPI (rev-phase), fMRI data was collected with reversed phase-encode blips, resulting in pairs of images with distortions going in opposite directions (5 volumes in total). From these pairs, the susceptibility-induced off-resonance field was estimated using a method similar to that described in Andersson, Skare (18) as implemented in FSL (19). The resulting voxel displacement map was used to warp individual volumes in the fMRI times series using the FSL TOPUP tool (https://fsl.fmrib.ox.ac.uk/fsl/fslwiki/TOPUP, accessed July 3rd, 2021). Preprocessing steps in SPM12, included slice time correction according to multi-slice acquisition, and spatial realignment to the mean volume to correct for rigid body movement in six directions. Mean functional images and corresponding aligned images were co-registered to navigator-based movement corrected and threshold T1 "robust" images derived from the MP2RAGE sequence. The individual T1 images were spatially normalized, and the resulting transformation was applied to the realigned and co-registered fMRI images, which were subsequently resampled to two mm isotropic resolution. Finally, resulting images were smoothed using a six mm full width at half maximum (FWHM) gaussian smoothing kernel. To minimize the effects of movement, fMRI image volumes with a frame-wise displacement (FD), a measure of the amount of movement of the head from one volume to the next (20), greater than one mm were removed together with one consecutive volume, by means of scan nulling regressors. Participants with more than 20% volumes excluded, were not included in the analyses (see **Inclusion of participants**).

**Statistical analyses**

*First level fMRI design*

This analysis included regressors that modelled congruent and incongruent events separately as main events of interest. Only events of correctly answered trials were included. The analysis further included the following separate regressors of no interest; left- and right-hand responses, errors of commission, errors of omission, trials eliciting a feedback stimulus, respiration, heartbeat, and realignment parameters, respectively. Parametric first-order polynomial regressors of interest were added to the main events to model RT-bin. Parametric second-order polynomial regressors of no interest were added to the main events to model congruency switches (i.e., events of congruent-to-incongruent and vice versa as an event of switch and congruent-to-congruent and incongruent-to-incongruent as events of no switch).

*Regressors of no interest*

Physiological measures, i.e., heart rate (3rd order expansion) and respiratory cycles (2nd order), recorded during the scanning session were included as nuisance regressors. The retrospective correction for noise in the fMRI time series was accomplished by use of the image based RETROICOR method (21), creating 10 regressors for implementation in the GLM. Rigid body realignment derived movement parameters (three translation and three rotation) as well as their temporal derivatives and corresponding squared regressors resulted in 24 additional nuisance regressors (22, 23) that were incorporated as regressors of no interest in the GLM. As SPM12 uses an effects of interest mask to estimate global AR(1) based pre-whitening parameters, these nuisance regressors were marked as effects of no interest during the restricted maximum likelihood procedure.

*Second level fMRI designs*

To examine group differences in brain activation during successful interference control, we used the contrast images of incongruent > congruent from the first level to conduct a one-way (group [FHR-SZ, FHR-BP, PBC] by successful interference) voxel-wise analysis of variance (ANOVA) in SPM12. We assessed the effect of successful interference with the mean contrast [0.333 0.333 .333] and group difference contrast $\begin{bmatrix} 1 & -1 & 0 \\ 0 & 1 & -1 \end{bmatrix}$.

To examine the linear relationship between brain activation and RT bin from the distributional analysis, we used the contrast images of incongruent > 0 and congruent > 0 from the first level to conduct a one-way (group [FHR-SZ, FHR-BP, PBC] by condition [congruent or incongruent]) voxel-wise ANOVA. We assessed the effect of linear relation to RT bin with the mean contrast [0.333 0.333 .333] and group difference contrast $\begin{bmatrix} 1 & -1 & 0 \\ 0 & 1 & -1 \end{bmatrix}$.

To examine the linear relationship between brain activation and reaction time variability ($RT_{CV}$), we used contrast images of incongruent > 0 and congruent > 0 from the first level to conduct a one-way (group [FHR-SZ, FHR-BP, PBC] by condition [congruent or incongruent]) voxel-wise ANOVA with $RT_{CV}$ added as a group-wise task-regressor of interest. We assessed the effect of linear relation to RT bin with the mean contrast [0.333 0.333 .333] and group difference contrast $\begin{bmatrix} 1 & -1 & 0 \\ 0 & 1 & -1 \end{bmatrix}$.

Areas of significant activation were identified using the probabilistic Harvard-Oxford Cortical and Subcortical Structural atlases, and the Juelich Histological Atlas (24), on the MNI152 2 mm resolution template image, implemented in FSLeyes. Areas were assigned according to highest probability.

**Region of interest analysis**

Creation of spheres and extraction of subject-specific mean parameter estimates from the nine regions of interest (ROIs) identified from the successful interference activation analyses was accomplished with the MarsBar (MARSeille Boîte À Région d'Intérêt, version 0.44 (25)) toolbox implemented in SPM12 and Matlab 2020a. We defined regions of interest (ROIs) as spheres with 10 mm radius centered in the coordinates according to the peak activation from the whole-brain successful interference activation analysis (eTable 2).

| Cluster forming threshold of $p < 0.001$ | | | | | |
|---|---|---|---|---|---|
| | Cluster level p-value (FWE corr.) | x (mm) | y (mm) | z (mm) | Harvard-Oxford atlas |
| **ROI I** | <.001 | -6 | -78 | -34 | 60% Left Crus II |
| **ROI II** | <.001 | 40 | -84 | -8 | 24% Right Lateral Occipital Cortex, inferior division |
| **ROI III** | <.001 | -36 | -86 | -6 | 49% Left Lateral Occipital Cortex, inferior division |
| **ROI IV** | | -30 | 22 | 4 | 63% Left Insular Cortex |
| **ROI V** | <.001 | 30 | 22 | 6 | 39% Right Insular Cortex |
| **ROI VI** | <.001 | 46 | 6 | 30 | 40% Right Precentral Gyrus |
| **ROI VII** | <.001 | -6 | 8 | 50 | 35% Left Juxtapositional Lobule Cortex (formerly supplemental Motor Cortex) |
| **ROI VIII** | <.001 | 28 | -2 | 52 | 25% Right Middle Frontal Gyrus |
| **ROI IX** | <.001 | -22 | -6 | 52 | 46% Left Superior Frontal Gyrus |

*eTable 2: Whole brain interference activation (incongruent trials > congruent trials) across groups at p < 0.001, uncorrected. The table states peak x, y, z coordinates used for region of interest (ROI) definition of the nine clusters. The Harvard-Oxford Cortical and Subcortical structural Atlases and the Cerebellar Atlas in MNI152 space, all implemented in FSLeyes, were used for identifying location of peak activation coordinates according to highest probability, after normalization of the study specific template to MNI152 space with FNIRT.*

*eTable 3: Whole-brain interference activation with time-bin as a first parametric modulator of interest across groups at p < 0.001, uncorrected. The table stat[illegible] wise error (FWE) corrected p-values and corresponding peak x, y, z coordinates of peak activation. The Harvard-Oxford Cortical and Subcortical structural Atl[illegible] Atlas in MNI152 space, all implemented in FSLeyes, were used for identifying location of peak activation coordinates according to highest probability, after no[illegible] specific template to MNI152 space with FNIRT.*

| Cluster-forming threshold of p < 0.001 | | | | | | | | | |
|---|---|---|---|---|---|---|---|---|---|
| Congruent > 0 | | | | | Incongruent > 0 | | | | |
| Cluster level p-value (FWE corr.) | x | y | z | Harvard-Oxford atlas | Cluster level p-value (FWE corr.) | x | y | z | Harvard-Oxford atlas |
| <.001 | -22 | 0 | 54 | 44% Superior frontal gyrus | <.001 | -2 | 10 | 48 | 61% Paracingulate Gyrus |
| <.001 | -58 | -30 | 44 | 62% Supramarginal Gyrus, anterior division | <.001 | -38 | -42 | 44 | 34% Superior Parietal Lobule |
| <.001 | -48 | 6 | 28 | 46% Precentral Gyrus | <.001 | -30 | 22 | 4 | 63% Insular Cortex |
| <.001 | -14 | -34 | -2 | 76% Left Thalamus | <.001 | 14 | -66 | 50 | 30% Lateral Occipital Cortex, superior division |
| 0.004 | 14 | -36 | 44 | 25% Precuneous Cortex | <.001 | 50 | -52 | 10 | 22% Middle Temporal Gyrus, temporooccipital part |
| <.001 | 50 | -52 | -16 | 64% Inferior Temporal Gyrus, temporooccipital part | <.001 | 4 | -20 | 0 | 84% Right Thalamus |
| .002 | -10 | 2 | 14 | 88% Left Caudate | <.001 | -16 | -72 | 6 | 44% Intracalcarine Cortex |
| <.001 | 50 | 8 | 18 | 35% Inferior Frontal Gyrus, pars opercularis | .026 | -10 | 2 | 12 | 88% Left Caudate |
| <.001 | 32 | 18 | 8 | 54% Insular Cortex | .037 | 20 | 2 | 4 | 57% Right Pallidum |
| .017 | -38 | 28 | 20 | 34% Inferior Frontal Gyrus, pars triangularis | | | | | |

| **Cluster-forming threshold of p < 0.001** | | | | |
|---|---|---|---|---|
| **Congruent > 0** | | | | |
| Cluster level p-value (FWE corr.) < 0.05 | x | y | z | Havard-Oxford atlas |
| .004 | 34 | -74 | -12 | 46% Right Occipital Fusiform Gyrus |
| .004 | 20 | -94 | 2 | 42% Right Occipital Pole |
| <.001 | -20 | -94 | -6 | 32% Left Occipital Pole |
| .017 | -26 | -44 | 44 | 22% Superior Parietal Lobule |

*eTable 4: Whole brain activation at congruent > 0 scales negatively with $RT_{CV}$ across groups at p < 0.001, uncorrected. The table states cluster-level, Family-wise error (FWE)-corrected p-values and corresponding peak x, y, z coordinates of peak activation. The Harvard-Oxford Cortical Atlas, implemented in FSL eyes, was used for identifying location of peak activation coordinates according to highest probability.*

| | FHR-BP | FHR-SZ | PBC | Condition effect | Group effect | Post hoc comparisons | | |
|---|---|---|---|---|---|---|---|---|
| | | | | | | FHR-BP vs FHR-SZ | FHR-SZ vs PBC | FHR-BP vs PBC |
| **Accuracy rate, %, mean (SD)** | | | | | | | | |
| Congruent | 98.4 (2.0) | 98.7 (2.3) | 99 (1.7) | $P$<.001[e] | $P$=.218[a] | - | - | - |
| Incongruent | 91.2 (7.6) | 93.7 (7.2) | 94.9 (6.0) | | $P$=.135[a] | - | - | - |
| **$\Delta_{accuracy}$, %, mean (SD)** | -7.2 (7.2) | -7.1 (6.1) | -5.6 (5.7) | - | $BF_{10}$=.26 | - | - | - |
| **Reaction time, ms, mean (SD)** | | | | | | | | |
| Congruent | 464.3 (56.9) | 465.1 (61.0) | 461.0 (57.2) | $BF_{10}$ > 100 | $BF_{10}$=.22 | - | - | - |
| Incongruent | 554.8 (62.7) | 552.3 (64.1) | 541.1 (57.3) | | | - | - | - |
| **ΔRT, ms, mean (SD)** | 90.5 (33.6) | 87.2 (35.4) | 80.0 (32.5) | - | $BF_{10}$=.32 | - | - | - |
| **$RT_{CV}$, %, mean (SD)** | | | | | | | | |
| Congruent | 24.6 (3.6) | 24.6 (4.7) | 23.3 (3.8) | $BF_{10}$ > 100 | $BF_{10}$=6.88 | $BF_{10}$=.13 | $BF_{10}$=8.04 | $BF_{10}$=24.31 |
| Incongruent | 20.1 (2.9) | 20.4 (3.1) | 18.7 (3.0) | | | | | |
| **$\Delta_{RTCV}$** | -4.5 (3.6) | -4.2 (4.2) | -4.6 (3.7) | - | $BF_{10}$=.08 | - | - | - |
| **Distributional analysis** | | | | | | | | |
| **Congruent accuracy rate, %, mean (SD)** | | | | | | | | |
| Timebin 1 | 97.8 (4.0) | 98.7 (3.9) | 9[illegible].0 ([illegible].8) | $P$=.034[b] | $P$=.126 | - | - | - |
| Timebin 2 | 99.1 (2.8) | 99.0 (2.8) | 9[illegible].4 ([illegible].9) | | $P$=.902 | - | - | - |
| Timebin 3 | 98.6 (3.3) | 98.9 (3.2) | 9[illegible].3 ([illegible].3) | | $P$=.268 | - | - | - |
| Timebin 4 | 99.0 (2.6) | 98.6 (3.0) | 98.4 ([illegible].6) | | $P$=.579 | - | - | - |
| **Incongruent accuracy rate, %, mean (SD)** | | | | | | | | |
| Timebin 1 | 81.3 (15.4) | 82.3 (15.5) | 83.[illegible] ([illegible].7) | $P$<.001[b] | $P$=.765 | - | - | - |
| Timebin 2 | 98.3 (3.3) | 97.7 (5.2) | 9[illegible].0 (3.0) | | $P$=.118 | - | - | - |
| Timebin 3 | 97.2 (5.2) | 96.9 (5.2) | 98.7 ([illegible].4) | | $P$=.020[c,d] | - | - | - |
| Timebin 4 | 95.0 (6.8) | 95.2 (6.3) | 96.8 ([illegible].4) | | $P$=.029[c,d] | - | - | - |
| **Dist-$\Delta_{resp\_acc}$, %, mean (SD)** | | | | | | | | |
| Timebin 1 | -16.5 (15.4) | -16.4 (14.2) | -15.[illegible] (11.9) | $BF_{10}$ > 100 | $BF_{10}$=.11 | - | - | - |
| Timebin 2 | -0.8 (3.5) | -1.4 (5.3) | -0.3 ([illegible].4) | | | - | - | - |
| Timebin 3 | -1.4 (6.5) | -2.0 (5.8) | -0.[illegible] (3.7) | | | - | - | - |
| Timebin 4 | -4.0 (7.4) | -3.4 (6.7) | -1.[illegible] ([illegible].2) | | | - | - | - |
| **Dist-$\Delta_{RT}$, ms, mean (SD)** | | | | | | | | |
| Timebin 1 | 76 (28) | 71 (29) | [illegible] ([illegible]) | $BF_{10}$ > 100 | $BF_{10}$=.15 | - | - | - |
| Timebin 2 | 89 (35) | 84 (32) | 78 ([illegible]) | | | - | - | - |
| Timebin 3 | 90 (44) | 87 (49) | 77 ([illegible]) | | | - | - | - |
| Timebin 4 | 88 (56) | 90 (68) | 79 ([illegible]) | | | - | - | - |

*eTable 5: Behavioral mean outcome variables and statistical inference from the behavioral analysis [illegible] schizophrenia (FHR-SZ), 63 children at familial high-risk of bipolar disorder (FHR-BP), and 98 population-[illegible] Danish High Risk and Resilience Study – VIA 11. Outcome variables were collected during an arrow versi[illegible] with functional magnetic resonance imaging (fMRI) at 3Tesla. SD; Standard deviation, $BF_{10}$; Levels of evide[illegible] interpretation of Bayes factors (BF) in favor of the alternative hypothesis ($BF_{10}$), Dist- $\Delta_{resp\text{-}acc}$; RT distribu[illegible] response accuracy, Dist- $\Delta_{RT}$; RT distributional analysis of the flanker effect on reaction time.*

[a] *Kruskal-Wallis test.*

[b] *Related samples Friedman's Two-Way analysis of variance (ANOVA).*

[c] *Independendt-Samples Kruskal-Wallis Test.*

[d] *post-hoc adjusting in according to Bonferroni correction with α = 0.05 results in non-significant P-values [illegible]*

[e] *Related samples Wilcoxon Signed Rank Test*

**A** Analysis of effects: ROI analysis of successful interference processing

| Effects | P(incl) | P(excl) | P(incl\|data) | P(excl\|data) | $BF_{incl}$ |
|---|---|---|---|---|---|
| **ROI** | 0.200 | 0.400 | 1.000 | 0.000 | ∞ |
| **Group** | 0.200 | 0.400 | 0.138 | 0.862 | 0.107 |
| **ROI x Group** | 0.200 | 0.400 | 9.38E-05 | 1.000 | 3.75E-04 |

**B** Analysis of effects: ROI analysis of congruent > 0

| Effects | P(incl) | P(excl) | P(incl\|data) | P(excl\|data) | $BF_{incl}$ |
|---|---|---|---|---|---|
| **ROI** | 0.600 | 0.400 | 1.000 | 5.11E-17 | 1.31E+14 |
| **Group** | 0.600 | 0.400 | 0.114 | 0.886 | 0.085 |
| **ROI x Group** | 0.200 | 0.800 | 4.23E-07 | 1 | 1.69E-06 |

**C** Analysis of effects: ROI analysis of incongruent > 0

| Effects | P(incl) | P(excl) | P(incl\|data) | P(excl\|data) | $BF_{incl}$ |
|---|---|---|---|---|---|
| **ROI** | 0.600 | 0.400 | 1.000 | 4.55E-15 | 1.47E+14 |
| **Group** | 0.600 | 0.400 | 0.052 | 0.948 | 0.036 |
| **ROI x Group** | 0.200 | 0.800 | 2.82E-06 | 1.000 | 1.13E-05 |

*eTable 6: Statistical output from the Bayesian repeated measures (RM) analyses of variance (ANOVA) with region of interest (ROI) as repeated measures with nine levels (ROI I: Left Crus II , ROI II: Right Lateral Occipital Cortex, ROI III: Left Lateral Occipital Cortex, ROI IV: Left Insular Cortex , ROI V: Right Insular Cortex, ROI VI: Right Precentral Gyrus , ROI VII: Left Juxtapositional Lobule Cortex , ROI VIII: Right Middle Frontal Gyrus, ROI IX: Left Superior Frontal Gyrus ) and Group as between subject factors with three levels (Familial high risk [FHR] of schizophrenia [SZ] n=85, FHR of bipolar disorder (BP) n=63, and population-based control [PBC] n=98) for* ***A)*** *the successful interference processing, and the interference activation with time-bin as a first parametric modulator of interest on* ***B)*** *congruent and* ***C)*** *incongruent trials. Levels of evidence are reported according to standard interpretation of Bayes factors (BF) in favor of the alternative hypothesis (BF10) ranging from 1 (no evidence) to 1-3 (anecdotal evidence), 3-10 (moderate evidence), 10-30 (strong evidence), 30-100 (very strong evidence), and >100 (decisive evidence). The BF quantifies the relative predictive performance of two rival hypotheses.*
